# Anatomical Predictors of Left Internal Carotid Artery Catheterization in Transradial Angiography

**DOI:** 10.1101/2021.10.07.21264684

**Authors:** Seon Woong Choi, Hoon Kim, Seong Rim Kim, Ik Seong Park, Sunghan Kim

## Abstract

**Introduction:** Transradial angiography (TRA) has received considerable attention in the field of neurointervention owing to its advantages over transfemoral approaches. However, the difficulty of left internal carotid artery (ICA) catheterization under certain anatomical conditions of the aortic arch and its branches is a limitation of TRA. This study aimed to investigate the anatomical predictors of successful catheterization of the left ICA in TRA.

**Materials and Methods:** From January 2020 to October 2020, 640 patients underwent TRA at a single institute. Among them, 263 consecutive patients who were evaluated by contrast-enhanced MRI before TRA were included in our study and assigned to success and failure groups, according to whether left ICA catheterization was possible or not. Anatomical predictors that may affect the success of left ICA catheterization in TRA were investigated for the purposes of our study.

**Results:** The multivariable analysis included variables that demonstrated significant univariate associations with ICA catherization (P<0.0001). Variables included in the model were the type of aortic arch, height of right subclavian artery, turn-off angle of the left common carotid artery (CCA), distance between innominate artery to the left CCA, angulation of right subclavian artery, and angulation of the left CCA, which we identified as significant predictors of left ICA catheterization.

**Conclusion:** Success of left ICA catheterization in TRA was related to the vascular geometry of the aortic arch and its branches. Evaluating the anatomical predictors identified in this study using pre-procedure imaging may enhance the success rate of left ICA catheterization in TRA.

## INTRODUCTION

Transradial angiography (TRA) has a number of advantages over transfemoral angiography for cerebral angiography, such as reduced rates of procedure-related complications and improved patient compliance, and TRA may additionally serve as a feasible, alternative route for patients who are pregnant or those with severe obesity and severe atherosclerotic disease of the ilio-femoral arteries.^1 2^ Despite these advantages, the main limitation of TRA is that the selective catheterization of the left internal carotid artery (ICA) and left vertebral artery (VA) may be more challenging in some patients.^3^ Selective left VA catheterization can be substituted by a technique involving placing the catheter tip in the left subclavian artery and injecting a contrast agent simultaneously, while inflating the blood pressure cuff on the left arm.^1^ However, selective left ICA catheterization is not possible under certain anatomical conditions of the aortic arch and its branches. Therefore, determining the factors that may affect the success of catheterization of the left ICA through the TRA is crucial for improving the adoption and utilization of TRA in the field of neurointerventional surgery. We aimed to investigate the clinical and anatomical predictors of a successful left ICA catheterization in TRA.

## MATERIALS AND METHODS

### Patient demographics

We performed a retrospective review of prospectively collected databases in a single institute between January 2020–October 2020. Among the patients who underwent TRA, those who were able to evaluate the state of the aortic arch and its branches by contrast-enhanced magnetic resonance angiography (CE-MRA), which was performed within 3 months of TRA, were selected as candidates for the study. The following cases were excluded from the study: 1) cases with any vasculopathy that can cause changes in normal vascular geometry such as moyamoya disease or over 50% of intracranial arterial stenosis; 2) cases with difficult navigation of the distal part of the left ICA, such as a history of carotid artery stenting or > 50% of proximal ICA stenosis; 3) if the navigation of branches of the aortic arch were unavailable with TRA due to anomalous branches of the aortic arch, such as the aberrant right subclavian artery (Arteria Lusoria)^4^; 4) if there was a problem with catheter manipulation due to complex radial artery anatomy such as radial loop; and 5) if vasospasm occurred during angiography, which interferes with the angiography procedure. In addition, referring to the results of previous studies that the presence of bovine aortic arch anatomy can ease the catheterization of the left ICA^1 5^, all cases with bovine arch were excluded for objective evaluation of other parameters affecting the left ICA catheterization. Finally, cases were assigned to the success and failure groups according to whether left ICA catheterization was possible or not. All clinical and radiological data were obtained from electronic medical records and a prospectively registered diagnostic TRA database.

### Radial angiography technique

The angiographic technique of TRA used in our study was similar to that used in previous studies.^2 3^ The right radial artery was used as an access channel for TRA in all patients in the study. A 5 Fr Glidesheath Slender (Terumo Interventional Systems, Terumo Medical Corporation North America, Somerset, NJ) 7 cm introducer sheath, a 5 Fr Radifocus Glidecath Simmons/Sidewinder type 2 catheter (Cook, Bloomington, IN, USA), and a 0.035-inch Radifocus guide wire (Terumo Interventional Systems, Terumo Medical Corporation Europe, Leuven, Belgium) were used as basic tools for TRA in all cases. If the left ICA catheterization was unsuccessful with basic tools, then a 5 Fr Radifocus Glidecath Simmons/Sidewinder type 3 catheter or Newton technique 4 catheter with or without a stiff type 0.035-inch Radifocus guide wire were used as rescue devices.

All angiography cases included in this study were conducted by neurosurgeons with at least 1,000 cases of TRA experience. If left ICA catheterization was unsuccessful despite the use of rescue devices, it was considered a failure of the left ICA catheterization.

### MRA imaging protocol

All patients underwent CE-MRA on an Achieva 3 Tesla magnet (Philips Medical Systems, Best, Netherlands) equipped with a SENSE NV 16-channel coil. The CE-MRA was performed after IV administration of 0.1 mmol/kg bodyweight gadobutrol (Gadovist®; Bayer Healthcare Pharmaceuticals, Berlin, Germany) with TR = 5.5 msec, TE = 1.52 msec, flip angle = 30°, FOV = 320×280 mm^2^, matrix = 516×302, reconstruction matrix = 1,024, slice thickness = 1.2, sensitivity encoding factor = 2 × 1, contrast bolus time = 23 sec, total acquisition time: contrast bolus time = 10:1.

### Anatomical evaluation and definition of predictors

Contrast-enhanced MRA data were transferred to a three-dimensional workstation and reconstructed for measurement of each anatomical predictor. Measurements of anatomical predictors were made by two independent certified neurosurgeons who were blinded to the patients’ information. If the measurements of the two neurosurgeons differed, a third neurosurgeon reviewed the data, and a final consensus was reached by the three investigators regarding the results.

The definition of each anatomical predictor is as follows: (1) The type of aortic arch was categorized into three according to Casserly’s classification.^6^; (2) The height of the right subclavicular artery was defined as the distance between the axial planes drawn at the topmost point of the right subclavicular artery and at the medial border of the origin of the innominate artery, respectively.; (3) Turn-off angle of left common carotid artery (CCA) was the angle between the centerline of the innominate artery and left CCA.^7^ Each centerline was drawn parallel to the direction of each artery from the origin until the vessel forming the first angulation.; (4) Distance between the innominate artery to the left CCA was defined as the distance along the upper border of aortic arch between the medial border of the innominate artery and medial border of the left CCA orifice.; (5) Diameter of the left CCA was measured at its widest point.; (6) Angulation of the right subclavian artery, left CCA, and left ICA was classified into three categories: tortuosity, kinking, and looping according to the modified criteria of Wiebel-Fields and Metz.^8 9^ Tortuosity was defined if S- or C-shaped elongation or undulation was noted in the course of each vessel. Angulation of the two segments of each vessel forming the kink measured at ≥ 60° was also considered tortuosity. Kinking was defined if the angulation of the two segments of each vessel was 30°–60°. Looping was defined as the elongation or redundancy of each vessel, resulting in an exaggerated S-shaped curve or a circular configuration. Angulation of the two segments of each vessel forming the kink measured < 30° was considered looping as well. The definitions of each anatomical predictor are summarized in Table 1.

**Table 1.** Definition of each anatomical predictor for left ICA catheterization

| Variables | Definition |
| --- | --- |
| Aortic arch anatomy | Classified into the three types according to the Casserly classification: Type 1, 2, 3 |
| Height of right subclavian artery | Distance between the axial planes drawn at the topmost point of the right subclavicular artery and at the medial border of the origin of the innominate artery |
| Turn-off angle of left CCA | Angle between the centerline of innominate artery and left CCA |
| Distance between innominate artery to left CCA | Distance along the upper border of aortic arch between the medial border of the innominate artery and medial border of left CCA orifice |
| Diameter of left CCA | Measured at the widest point of left CCA |
| Angulation of right subclavian artery, left CCA & ICA | Classified into the three categories according to the modified criteria of Wiebel-Fields and Metz: tortuosity, kinking, and looping<br>Angulation was defined as angle between vessel segment: Tortuosity $\geq 60^\circ$, kinking $30^\circ$–$60^\circ$, looping $< 30^\circ$ |

ICA, internal carotid artery; CCA, common carotid artery.

### Statistical analysis

Continuous variables were described as mean ± standard deviation, and categorical variables were summarized as frequencies and percentages (%). A Student’s t-test was used for analyses of continuous variables. The Pearson χ^2^ test or Fisher’s exact test was used to analyze the categorical variables. Univariate logistic regression analysis was performed to determine the association between the demographic and anatomical predictors associated with left ICA catheterization. Multivariable logistic regression analysis was performed for variables with an adjusted effect, and a P < 0.05, in univariate analysis, was used to determine associations between the parameters and left ICA catheterization (multivariable analysis 1). To prevent the logistic regression model from overfitting the data, additional multivariable logistic regression analysis was performed for variables with an adjusted effect, and P < 0.0001 in univariate analysis (multivariable analysis 2). Results of binary logistic regression were reported as odds ratios (ORs) with a 95% confidence interval (CI). Statistical significance was set at P < 0.05. Firth’s bias correction method was used to reduce small-sample bias due to separation in logistic regression with rare events.^10^ The threshold value for the cut-off point of the continuous variable among the anatomical predictors for the catheterization of the left ICA was determined using receiver operating characteristic (ROC) analysis and Youden’s index. Binary logistic regression was performed on the possibility of left ICA catheterization based on the cut-off point. The diagnostic performance of each binomial variable was assessed. All statistical analyses were performed using SAS version 9.4 (SAS Inc., Cary, NC, USA).

## Results

### Patient Demographics

Subjects in the failure group were older and showed higher prevalence of hypertension, diabetes mellitus, coronary artery disease, and ischemic stroke.

In terms of anatomical predictors, the success and failure groups showed significant differences in all anatomical variables. The predominant aortic arch form was type 1 in the success group, while the type 3 arch was predominant in the failure group. In addition, the failure group had significantly higher values of height of the right subclavian artery, lower values of turn-off angle of the left CCA, higher values of distance between the innominate artery and left CCA, and diameter of the left CCA. Angulation of the right subclavian artery, left CCA, and left ICA was significantly more severe in the failure group. The demographic characteristics of the two groups are presented in Table 2.

**Table 2.** Comparison of baseline characteristics between success group and failure group

| Variables | Success group (n = 146) | Failure group (n = 117) | p value |
| --- | --- | --- | --- |
| Age (years) | 56.3±11.4 | 70.1±9.3 | <.0001 |
| Female | 85 (58.2) | 65 (55.6) | 0.6645 |
| Risk factors |  |  |  |
| Hypertension | 54 (37) | 90 (76.9) | <.0001 |
| Diabetes mellitus | 33 (22.6) | 42 (35.9) | 0.0176 |
| Dyslipidemia | 26 (17.8) | 19 (16.2) | 0.7371 |
| Coronary artery disease | 4 (2.7) | 13 (11.1) | 0.0061 |
| Ischemic stroke | 26 (17.8) | 44 (37.6) | 0.0003 |
| Smoking | 33 (22.6) | 22 (18.8) | 0.4515 |
| Type of aortic arch |  |  | <.0001 |
| Type 1 | 69 (47.3) | 11 (9.4) |  |
| Type 2 | 59 (40.4) | 43 (36.8) |  |
| Type 3 | 18 (12.3) | 63 (53.8) |  |
| Height of right subclavian artery (mm) | 50.1±9.7 | 58.6±13.1 | <.0001 |
| Turn-off angle of left CCA (°) | 19.8±13.9 | 9.1±10.1 | <.0001 |
| Distance between innominate artery to left CCA (mm) | 3.4±1.5 | 6.1±3.1 | <.0001 |
| Diameter of left CCA (mm) | 5.9±0.9 | 6.7±1.1 | <.0001 |
| Angulation of right subclavian artery |  |  | <.0001* |
| Normal | 0 (0) | 0 (0) |  |
| Tortous | 99 (67.8) | 29 (24.8) |  |
| Kinking | 47 (32.2) | 81 (69.2) |  |
| Looping | 0 (0) | 7 (6) |  |
| Angulation of left CCA |  |  | <.0001* |
| Normal | 54 (37) | 3 (2.6) |  |
| Tortous | 86 (58.9) | 63 (53.8) |  |
| Kinking | 6 (4.1) | 51 (44.6) |  |
| Looping | 0 (0) | 0 (0) |  |
| Angulation of left ICA |  |  | <.0001* |
| Normal | 3 (2.1) | 0 (0) |  |
| Tortous | 103 (70.5) | 57 (48.7) |  |
| Kinking | 40 (27.4) | 51 (43.6) |  |
| Looping | 0 (0) | 9 (7.7) |  |

Values are presented as mean ± standard deviation or number (%) as otherwise indicated.
\* p value using Fisher's exact test.
CCA, common carotid artery; ICA, internal carotid artery

### Clinical and Anatomical Predictors for Left ICA catheterization

The association of different clinical and anatomical predictors with left ICA catheterization is shown in Table 3. Univariate analysis revealed that old age, history of hypertension, diabetes mellitus, coronary artery disease, and ischemic stroke were associated with successful left ICA catheterization. Regarding anatomical predictors, a higher degree of aortic arch type, higher height of the right subclavian artery, narrower turn-off angle of the left CCA, wider distance between the innominate artery and left CCA, and larger left CCA diameter were significant clinical predictors for left ICA catheterization. Angulation of the right subclavian artery and left CCA were significantly associated with catheterization of the left ICA. In terms of angulation of the left ICA, the looping of the left ICA was associated with left ICA catheterization.

**Table 3.**
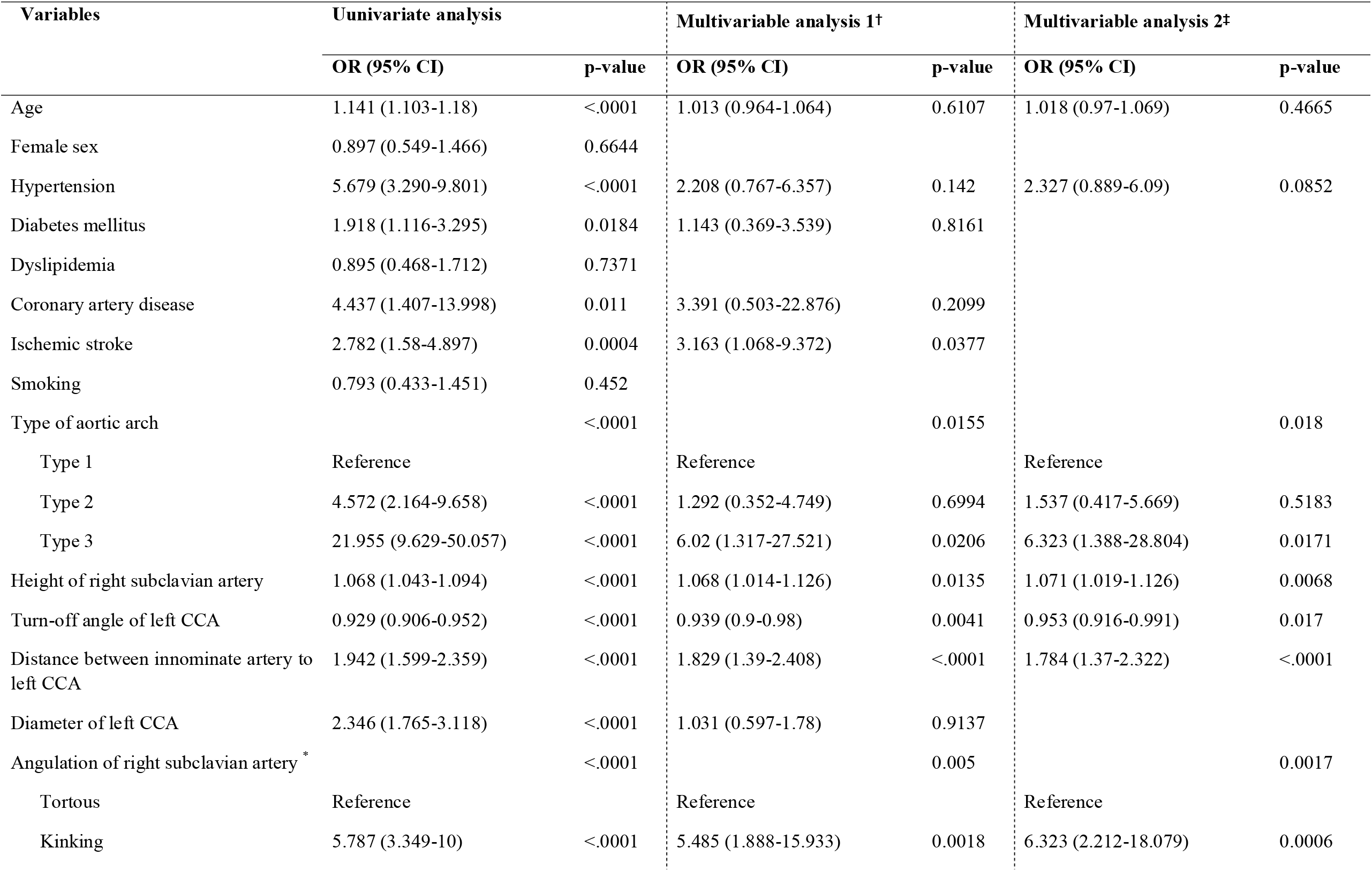
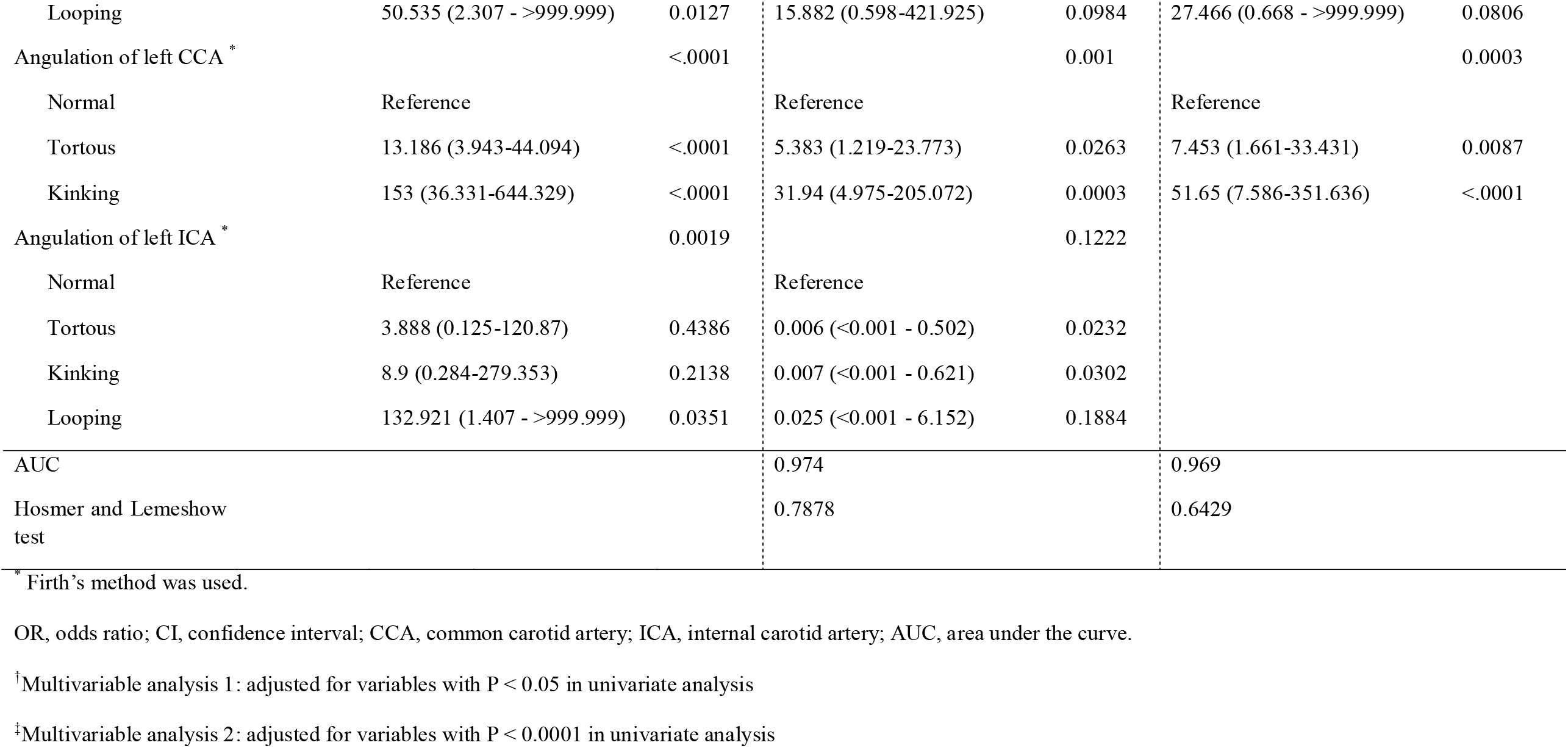
Univariate and multivariable analysis of clinical and anatomical predictors for left ICA catheterization.

| Variables | Univariate analysis |  | Multivariable analysis 1<sup>†</sup> |  | Multivariable analysis 2<sup>‡</sup> |  |
| --- | --- | --- | --- | --- | --- | --- |
|  | OR (95% CI) | p-value | OR (95% CI) | p-value | OR (95% CI) | p-value |
| Age | 1.141 (1.103-1.18) | <.0001 | 1.013 (0.964-1.064) | 0.6107 | 1.018 (0.97-1.069) | 0.4665 |
| Female sex | 0.897 (0.549-1.466) | 0.6644 |  |  |  |  |
| Hypertension | 5.679 (3.290-9.801) | <.0001 | 2.208 (0.767-6.357) | 0.142 | 2.327 (0.889-6.09) | 0.0852 |
| Diabetes mellitus | 1.918 (1.116-3.295) | 0.0184 | 1.143 (0.369-3.539) | 0.8161 |  |  |
| Dyslipidemia | 0.895 (0.468-1.712) | 0.7371 |  |  |  |  |
| Coronary artery disease | 4.437 (1.407-13.998) | 0.011 | 3.391 (0.503-22.876) | 0.2099 |  |  |
| Ischemic stroke | 2.782 (1.58-4.897) | 0.0004 | 3.163 (1.068-9.372) | 0.0377 |  |  |
| Smoking | 0.793 (0.433-1.451) | 0.452 |  |  |  |  |
| Type of aortic arch |  | <.0001 |  | 0.0155 |  | 0.018 |
| Type 1 | Reference |  | Reference |  | Reference |  |
| Type 2 | 4.572 (2.164-9.658) | <.0001 | 1.292 (0.352-4.749) | 0.6994 | 1.537 (0.417-5.669) | 0.5183 |
| Type 3 | 21.955 (9.629-50.057) | <.0001 | 6.02 (1.317-27.521) | 0.0206 | 6.323 (1.388-28.804) | 0.0171 |
| Height of right subclavian artery | 1.068 (1.043-1.094) | <.0001 | 1.068 (1.014-1.126) | 0.0135 | 1.071 (1.019-1.126) | 0.0068 |
| Turn-off angle of left CCA | 0.929 (0.906-0.952) | <.0001 | 0.939 (0.9-0.98) | 0.0041 | 0.953 (0.916-0.991) | 0.017 |
| Distance between innominate artery to left CCA | 1.942 (1.599-2.359) | <.0001 | 1.829 (1.39-2.408) | <.0001 | 1.784 (1.37-2.322) | <.0001 |
| Diameter of left CCA | 2.346 (1.765-3.118) | <.0001 | 1.031 (0.597-1.78) | 0.9137 |  |  |
| Angulation of right subclavian artery * |  | <.0001 |  | 0.005 |  | 0.0017 |
| Tortous | Reference |  | Reference |  | Reference |  |
| Kinking | 5.787 (3.349-10) | <.0001 | 5.485 (1.888-15.933) | 0.0018 | 6.323 (2.212-18.079) | 0.0006 |
| Looping | 50.535 (2.307 - >999.999) | 0.0127 | 15.882 (0.598-421.925) | 0.0984 | 27.466 (0.668 - >999.999) | 0.0806 |
| Angulation of left CCA * |  | <.0001 |  | 0.001 |  | 0.0003 |
| Normal | Reference |  | Reference |  | Reference |  |
| Tortous | 13.186 (3.943-44.094) | <.0001 | 5.383 (1.219-23.773) | 0.0263 | 7.453 (1.661-33.431) | 0.0087 |
| Kinking | 153 (36.331-644.329) | <.0001 | 31.94 (4.975-205.072) | 0.0003 | 51.65 (7.586-351.636) | <.0001 |
| Angulation of left ICA * |  | 0.0019 |  | 0.1222 |  |  |
| Normal | Reference |  | Reference |  |  |  |
| Tortous | 3.888 (0.125-120.87) | 0.4386 | 0.006 (<0.001 - 0.502) | 0.0232 |  |  |
| Kinking | 8.9 (0.284-279.353) | 0.2138 | 0.007 (<0.001 - 0.621) | 0.0302 |  |  |
| Looping | 132.921 (1.407 - >999.999) | 0.0351 | 0.025 (<0.001 - 6.152) | 0.1884 |  |  |
| AUC |  |  | 0.974 |  | 0.969 |  |
| Hosmer and Lemeshow test |  |  | 0.7878 |  | 0.6429 |  |

\* Firth's method was used.
OR, odds ratio; CI, confidence interval; CCA, common carotid artery; ICA, internal carotid artery; AUC, area under the curve.
<sup>†</sup>Multivariable analysis 1: adjusted for variables with P < 0.05 in univariate analysis
<sup>‡</sup>Multivariable analysis 2: adjusted for variables with P < 0.0001 in univariate analysis

Multivariable analysis 1 revealed that the following were significant predictors of left ICA catheterization: history of ischemic stroke, aortic arch type, height of the right subclavian artery, turn-off angle of the left CCA, distance between innominate artery to the left CCA, angulation of the right subclavian artery, and angulation of the left CCA. All anatomical factors except angulation of the left ICA and diameter of the left CCA were included in the multivariable analysis 2 and all of them were also found to be significant predictors of left ICA catheterization: type of aortic arch, height of the right subclavian artery, turn-off angle of the left CCA, distance between the innominate artery to the left CCA, angulation of the right subclavian artery, and angulation of the left CCA. The area under the ROC curve (AUC) of multivariable analyses 1 and 2 were 0.974 and 0.969, respectively.

### Cut-Off Points of Anatomical Predictors for Left ICA catheterization

ROC curves were drawn for the height of the right subclavian artery, distance between the innominate artery to the left CCA, turn-off angle of the left CCA, and diameter of the left CCA. The ROC curve for the height of the right subclavian artery showed an AUC of 0.679. The cut-off value of the height of the right subclavian artery was determined to be 54.83 mm, for which the sensitivity and specificity were 0.633 and 0.726, respectively. The ROC curve for the turn-off angle of the left CCA showed an AUC of 0.684. The cut-off value of the turn-off angle of the left CCA was determined to be 17°, for which the sensitivity and specificity were 0.855 and 0.514, respectively. The ROC curve for the distance between the innominate artery and the left CCA showed an AUC of 0.745. The cut-off value of the distance between the innominate artery and left CCA was determined to be 4.25 mm, for which the sensitivity and specificity were 0.709 and 0.781, respectively. The ROC curve for the diameter of the left CCA showed an AUC of 0.671. The cut-off value of the diameter of the left CCA was determined to be 6.05 mm, for which the sensitivity and specificity were 0.718 and 0.623, respectively. The results are summarized in Table 4.

**Table 4.** Cut-off points of continuous variables to success of left ICA catheterization and its diagnostic performance.

| Variables | AUC (95% CI) | Cut-off point | Sensitivity (95% CI) | Specificity (95% CI) | OR (95% CI) | p-value |
| --- | --- | --- | --- | --- | --- | --- |
| Height of right subclavian artery | 0.679 (0.622-0.736) | $\geq 54.83$ | 0.633 (0.545-0.720) | 0.726 (0.654-0.798) | 4.56 (2.703-7.694) | <.0001 |
| Turn-off angle of left CCA | 0.684 (0.632-0.736) | $< 17$ | 0.855 (0.791-0.919) | 0.514 (0.433-0.595) | 6.213 (3.383-11.413) | <.0001 |
| Distance between innominate artery to left CCA | 0.745 (0.692-0.798) | $\geq 4.25$ | 0.709 (0.627-0.792) | 0.781 (0.714-0.848) | 8.697 (4.97-15.217) | <.0001 |
| Diameter of left CCA | 0.671 (0.614-0.728) | $\geq 6.05$ | 0.718 (0.636-0.800) | 0.623 (0.545-0.702) | 4.212 (2.495-7.11) | <.0001 |

OR, odds ratio; CI, confidence interval; AUC, area under the curve; CCA, common carotid artery.

## DISCUSSION

Our aim was to evaluate the anatomical features of the aortic arch and its branches that correlated with left ICA catheterization in the right TRA. The results of this study indicated that the type of aortic arch, height of the right subclavian artery, distance between the innominate artery and left CCA, turn-off angle between the innominate artery and left CCA, angulation of the right subclavian artery, and angulation of the left CCA were the anatomical predictors that affected left ICA catheterization.

The clinical benefits of TRA have generated interest in adopting the transradial approach for neuroendovascular procedures,^11^ but neurointerventionalists should become familiar with new techniques to perform TRA proficiently. One of the most important techniques is reforming the loop of the Simmons catheter successfully and maintaining it in a stable position to navigate the aortic arch and selectively catheterize the cerebral vessels.^12^ For selective catheterization of the left ICA using the Simmons catheter in the TRA, it is important to place the loop of the Simmons catheter in the trunk of the CCA, while maintaining a stable configuration. If the distal tip of the Simmons catheter advanced enough through the CCA, the loop of the Simmons catheter can become straightened and provide proximal support for the guide wire to navigate the targeted vessel (Figure 1). However, if the position of the catheter is unstable and the loop is not fully unfolded, the catheter will be herniated back into the aortic arch when attempting to advance the guide wire into the left ICA for navigation (Figure 2, 3). All anatomical predictors that affect the left ICA catheterization investigated in this study were related to the proximal support stability of the Simmons loop.

**Figure 1.**
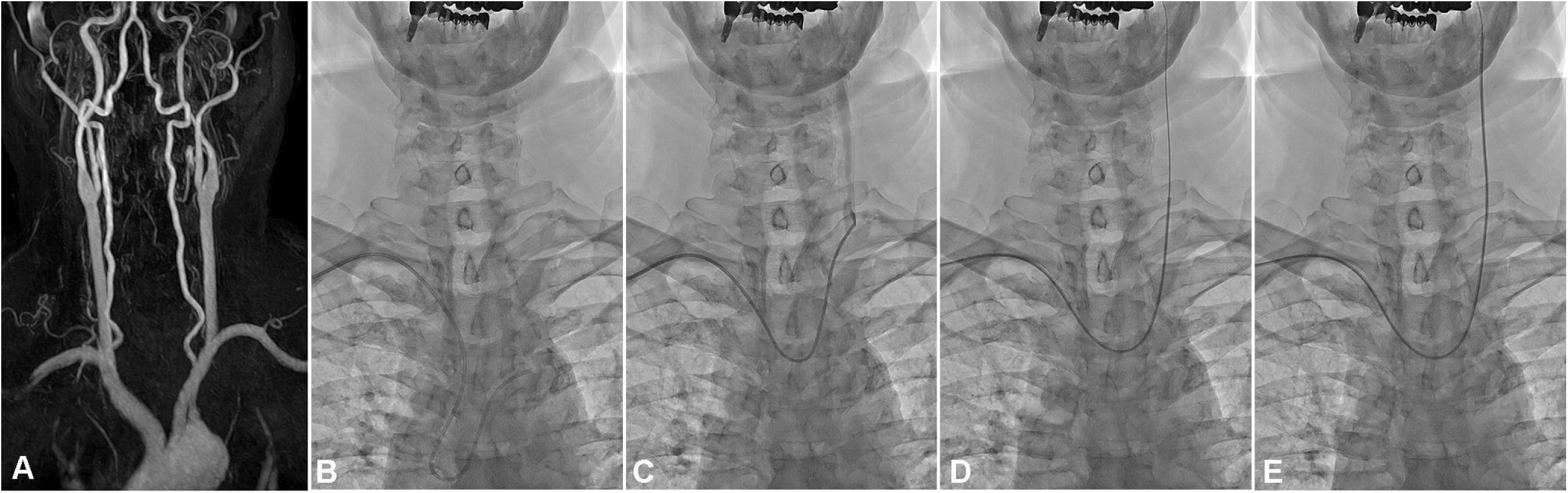
Success of left ICA catheterization during TRA. (A) Type 1 aortic arch with minimal angulation of the innominate artery and left CCA. (B) The loop of Simmons catheter is located in the ascending aortic arch. (C) The loop of Simmons catheter is placed in the trunk of the CCA with a stable configuration. (D, E) The guide wire and Simmons catheter can navigate the left ICA with proximal support from the loop of the Simmons catheter. ICA, internal carotid artery; TRA, transradial angiography; CCA, common carotid artery.

**Figure 2.**
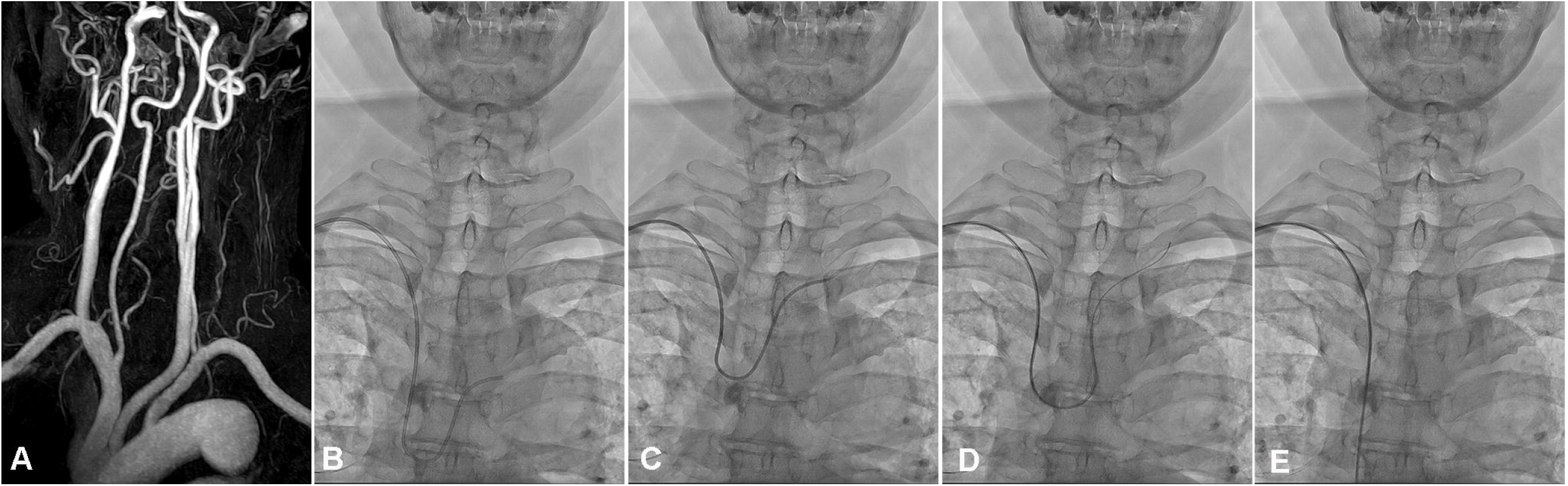
Failure of left ICA catheterization because of the elongation of the ascending aortic arch. (A) Type 3 aortic arch with high height of right subclavian artery and narrow turn-off angle between the innominate artery and left CCA. (B, C) Position of the loop of the Simmons catheter is unstable and the loop cannot be fully unfolded. (D, E) When the guide wire tries to advance into the left ICA, the loop of the Simmons catheter is herniated back into the aortic arch because an unstable loop of Simmons cannot provide enough proximal support for the navigation of the left ICA. ICA, internal carotid artery; CCA, common carotid artery

**Figure 3.**
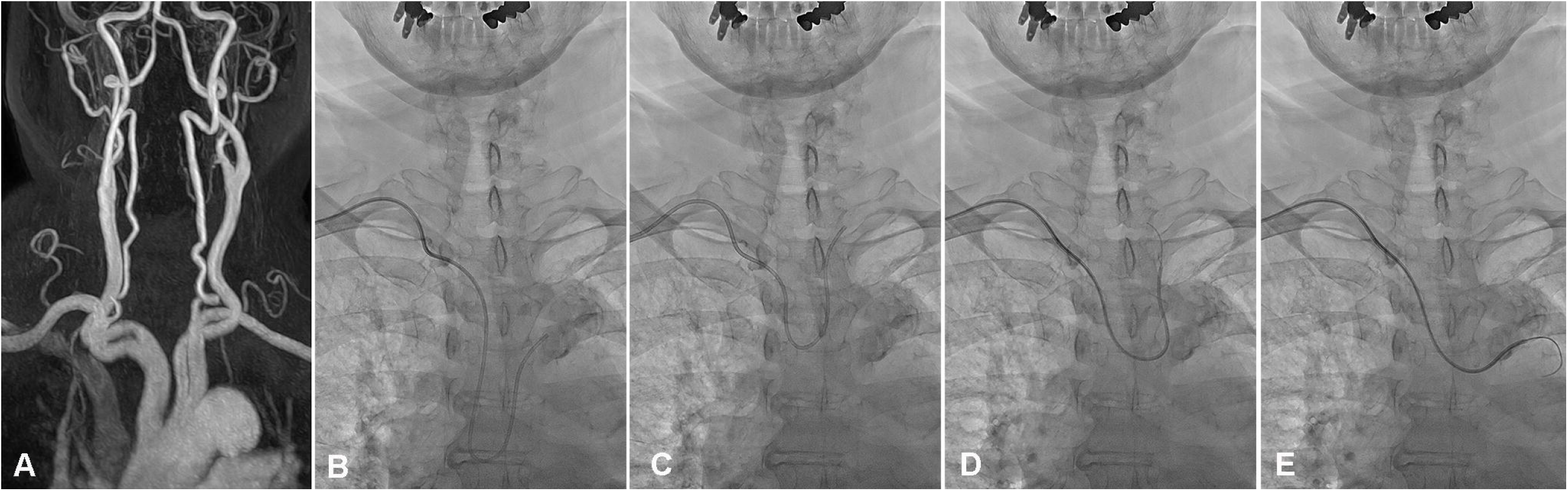
Failure of left ICA catheterization because of the angulation of the right subclavian artery and the left CCA. (A) Type 2 aortic arch with the kinking of the right subclavian artery and the left CCA. (B, C) The distal tip of the Simmons catheter cannot advance enough into the CCA because of severe angulation of the proximal portion of the left CCA. (D, E) When the guide wire tries to advance into the left ICA, the loop of the Simmons catheter is herniated back into the aortic arch because the unstable loop of Simmons cannot sufficiently support the proximal portion of the guide wire to overcome the loading of the catheter due to kinking of the left CCA. ICA, internal carotid artery; CCA, common carotid artery

In this study, the type of aortic arch and the height of the right subclavian artery were associated with catheterization of the left ICA during TRA. The type of aortic arch, height of the right subclavian artery, and the left CCA turn-off angle are all interrelated anatomical factors that are induced by the elongation of the aortic arch. In the type I aortic arch, the vertical distance between the orifice of the brachiocephalic artery and that of the left subclavian artery is fairly short. However, in the type II or III aortic arch, this distance is rather long. In addition, as the aortic arch type progresses from 1 to 3, the ascending aorta approximates a perpendicular position because of the elongation of the aortic arch, making the innominate artery and the left CCA parallel, thereby narrowing the left CCA turn-off angle. These anatomical changes can produce an acute curve at the point where the Simmons diagnostic catheter enters the left CCA.^5^ These anatomical features can prevent the Simmons loop from sufficiently entering the left CCA to make proper proximal support, which causes kickbacks of the catheter when the guide wire tries to navigate the Lt ICA (Figure 2). Changes in the distance between the innominate artery and left CCA are also related to elongation of the aortic arch. As the aortic arch elongated, the distance between the origin of the innominate artery and left CCA increased. The gap between the innominate artery and the left CCA makes the Simmons catheter unable to maintain a stable configuration for sufficient proximal support while accessing the left ICA.

This study corroborated that the angulation of the right subclavian artery and left CCA can affect the catheterization of the left ICA in TRA. Unlike the transfemoral approach, which accesses the target arteries through a relatively large and straight-running descending aorta, the angulation of a relatively narrow and three-dimensional driving right subclavian artery can affect the success of TRA. Angulation of the right subclavian artery can reduce the torquability and trackability of the catheter, which interferes not only with left ICA catheterization but also with the manipulation of the catheter in the TRA.^5^ The angulation of the left CCA is associated with the stability of the Simmons loop. Kinking or looping in the proximal CCA makes it difficult for the Simmons catheter to form a stable configuration of the loop. Moreover, distortion of the proximal CCA can act as a geometrical obstacle to the navigation of the guide wire, which induces herniation of the loop by placing more load on the catheter (Figure 3).

Several anatomical predictors were found to be less correlated with left ICA catheterization. Unlike the angulation of the left CCA, angulation of the left ICA was not associated with left ICA catheterization in TRA in this study. This may occur because the angulation of the left ICA does not directly affect the configuration of stability of the Simmons loop. Angulation of the left ICA may not cause a load to interfere with the guide wire navigating the left ICA. In addition, the diameter of the left CCA did not affect left ICA catheterization for TRA in this study. Because the stability and trackability of the catheter during angiography are closely related to the size of the navigating vessel, we hypothesized that if the diameter of the CCA becomes larger, it might induce instability of the catheter. However, there was no significant relationship between the diameter of CCA and left ICA catheterization in this study, which is presumably due to the small difference between the maximum and minimum values of the CCA diameter.

This study has limitations. First, it was designed retrospectively and included a small number of subjects from a single center. Second, the technical limitations of the TRA procedure may have affected the results of this study. Even though all TRA cases included in this study were performed by highly experienced neurosurgeons who performed TRA in over 1,000 cases, the possibility of left ICA catheterization may change depending on the dexterity and experience of the operators, which can be a potential source of bias. Third, device selection could also be a cause of bias. Catheter angiography is a device-dependent procedure, and the success rate of the procedure can be changed by selecting the device. All cases of TRA included in this study were performed using the Simmons catheter and 0.035-inch guide wire, which are most commonly used in TRA. However, catheters and wires of various diameters and shapes are available on the market, and the success rate of left ICA catheterization may be increased by combining these instruments. Fourth, the CE-MRA used for the evaluation of anatomical predictors in this study may not fully identify the anatomical characteristics of individual patients. Efforts were made to measure all anatomical predictors as close to reality as possible, but differences between actual and measured values may have occurred because of the three-dimensional shape of the aortic arch and its branches. Despite these limitations, the results of this study may contribute to increasing the utilization of TRA by presenting the criteria for left ICA catheterization in TRA.

## CONCLUSION

Successful left ICA catheterization during TRA was related to the vascular geometry of the aortic arch and its branches. The aortic arch type, height of the right subclavian artery, distance between the innominate artery and left CCA, turn-off angle between the innominate artery and left CCA, angulation of the right subclavian artery, and angulation of the left CCA are anatomical predictors that affect left ICA catheterization. Evaluating the anatomical predictors identified in this study using pre-procedure imaging may enhance the success rate of left ICA catheterization in TRA.

## Data Availability

The relevant anonymized patient level data are available on reasonable request from the authors.

## Acknowledgments

The authors thank Hye Rim Kim from Biostatistics Collaboration Unit, Department of Biomedical Systems Informatics, Yonsei University College of Medicine for statistical analysis.

## Contributors

All authors have made significant contributions to this work. All have read and approved the final version of this manuscript.

## Patient consent

Not required.

## Ethics approval

This Study was approved by the institutional review board of Bucheon St. Mary’s hospital, and the requirement for informed consent was waived. (subject number: HC21RISI0041) All procedures performed in the studies involving human participants were in accordance with the ethical standards of our Institutional Review Board with the 1964 Helsinki Declaration and its later amendments or comparable ethical standards. In this retrospective study, the requirement for informed consent was waived. Patient data was de-identified data upon data collection.

## Data availability statement

The relevant anonymized patient level data are available on reasonable request from the authors.

